# Rapid and extraction-free detection of SARS-CoV-2 from saliva with colorimetric LAMP

**DOI:** 10.1101/2020.05.07.20093542

**Authors:** Matthew A. Lalli, S. Joshua Langmade, Xuhua Chen, Catrina C. Fronick, Christopher S. Sawyer, Lauren C. Burcea, Michael N. Wilkinson, Robert S. Fulton, Michael Heinz, William J. Buchser, Richard D. Head, Robi D. Mitra, Jeffrey Milbrandt

## Abstract

Rapid, reliable, and widespread testing is required to curtail the ongoing COVID-19 pandemic. Current gold standard nucleic acid tests are hampered by supply shortages in critical reagents including nasal swabs, RNA extraction kits, personal protective equipment (PPE), instrumentation, and labor. Here we present an approach to overcome these challenges with the development of a rapid colorimetric assay using reverse-transcription loop-mediated isothermal amplification (RT-LAMP) optimized on human saliva samples without an RNA purification step. We describe our optimizations of the LAMP reaction and saliva pretreatment protocols that enabled rapid and sensitive detection of < 10^2^ viral genomes per reaction in contrived saliva controls. Moreover, our saliva pretreatment protocol enabled sensitive viral detection by conventional quantitative reverse transcription polymerase chain reaction (qRT-PCR) without RNA extraction. We validated the high performance of these assays on clinical samples and demonstrate a promising approach to overcome the current bottlenecks limiting widespread testing.

## Introduction

Establishing rapid and widespread testing for coronavirus disease 2019 (COVID-19) is essential to containing the pandemic and safely reopening society. The current gold standard test measures viral nucleic acids extracted from clinical swabs by quantitative reverse transcription polymerase chain reaction (qRT-PCR). This assay requires trained medical personnel, specialized instrumentation, supply-limited reagents, and significant technical labor. Isothermal nucleic acid amplification tests are an alternative to conventional PCR methods that do not require expensive instruments or trained personnel to perform the reaction or interpret the results. Specifically, loop-mediated isothermal amplification (LAMP) with simultaneous reverse-transcription (RT-LAMP) allows for rapid and sensitive detection of nucleic acids within one hour in an easily interpretable colorimetric assay that requires only a heat source^1,2^.

Several groups around the world are currently developing LAMP-based protocols for the detection of severe acute respiratory syndrome coronavirus 2 (SARS-CoV-2), the virus causing COVID-19^3-7^. The sensitivity of LAMP on purified RNA compares well to qRT-PCR, and LAMP may achieve higher sensitivity on crude clinical samples^5^. The robustness of the LAMP *Bst* polymerase to PCR inhibitors makes it especially well-suited and widely used for pathogen detection in unpurified samples^8^. This confers a major potential advantage over current testing protocols as it enables skipping the cost-, labor-, time-, and reagent-consuming step of RNA extraction.

Saliva is a promising sample for expanding and facilitating testing due to the ease, safety, and non-invasive nature of its collection and its relatively high viral load^9,10^. Recognizing these benefits, the US Food and Drug Administration (FDA) has approved saliva collection and preservation devices for downstream COVID-19 testing. Direct comparison of saliva to nasopharyngeal (NP) swabs from the same individuals revealed that saliva samples provided more consistent and sensitive results for SARS-CoV-2 detection^11^. Most saliva-based methods, however, still employ RNA extraction followed by qRT-PCR.

Here, we sought to establish and optimize a simple LAMP-based assay for the qualitative detection of SARS-CoV-2 directly from saliva without an RNA extraction step.

## Results

### LAMP Primer Screening

To develop our assay, we first compared the performance of five sets of recently developed LAMP primer sets targeting different regions of the SARS-CoV-2 genome^3-6^. We used a commercially available colorimetric enzyme mix from New England Biolabs (NEB) to perform LAMP reactions on quantitative *in vitro* transcribed RNA standards corresponding to regions targeted by LAMP primers^12^. Of these, the NEB Gene N-A^3^ and Lamb *et* al.^4^ primers targeting the nucleocapsid (Gene N) and Orf1ab regions respectively had the highest sensitivity and lowest rates of false positives in water-only controls (Supplementary Figure 1). Demonstrating specificity to SARS-CoV-2, these primers had no cross-reactivity with MERS coronavirus controls. These primer sets were prioritized for further testing.

### Reaction Optimization in Simulated Samples

Next, we validated these primer sets on both RNA standards and heat-inactivated viral particles spiked into water or human saliva to simulate clinical samples (Figure 1A). Across both sets of LAMP primers, and for both RNA and particles, saliva strongly inhibited LAMP detection of SARS-CoV-2 compared to water (Fig. 1B). Particles were weakly and inconsistently detected in saliva whereas their detection in water was on par with detection of RNA (Fig. 1C) indicating the presence of an inhibitor in saliva that impairs the assay. We observed time sensitivity of the colorimetric assay especially when testing saliva samples. Many samples, including negative controls, turned yellow in LAMP reactions longer than 40 minutes due to non-specific amplification. We found that a 30-minute incubation provided a reliable readout.

**Figure 1:**
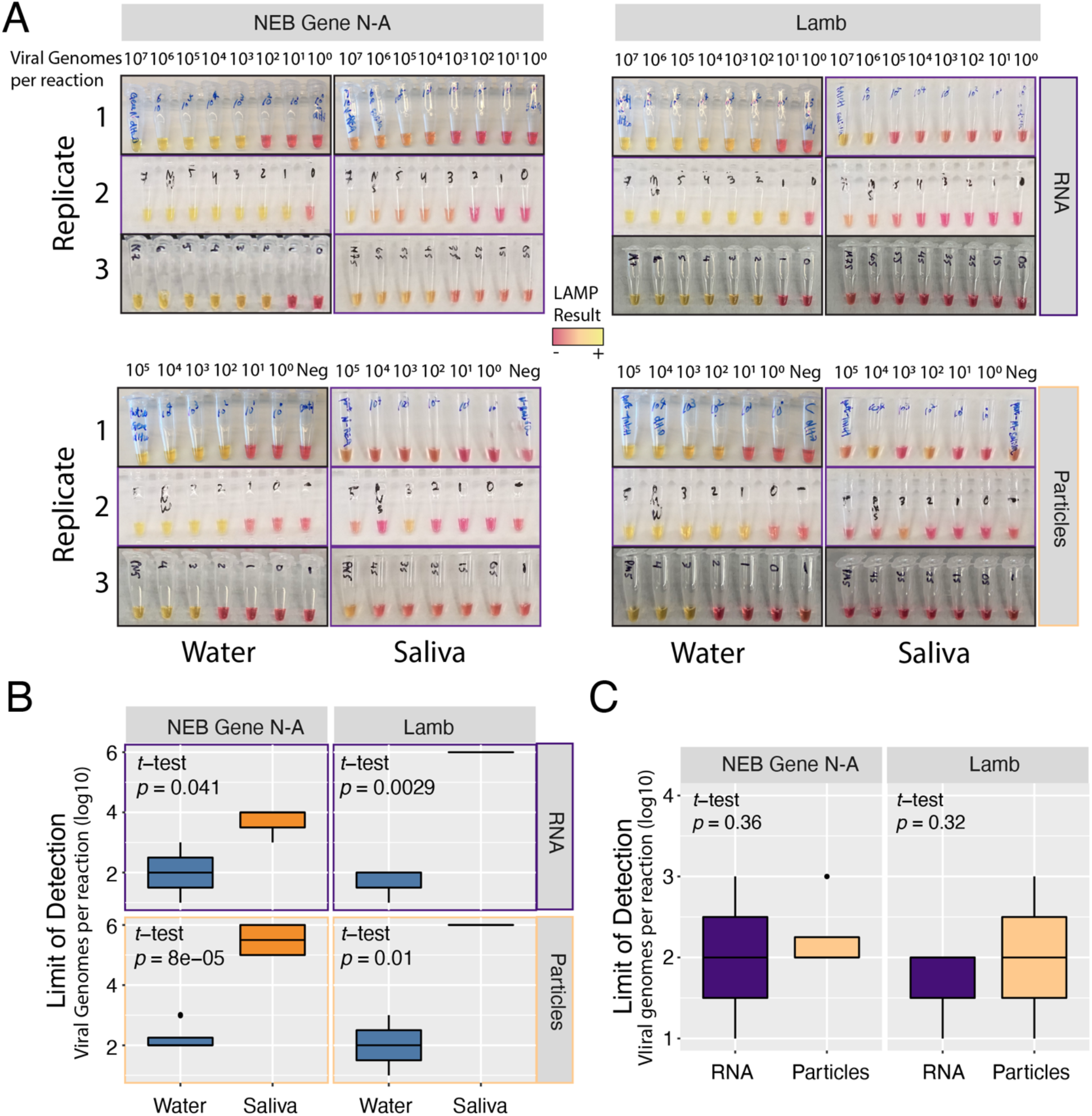
Assessing the sensitivity of detecting SARS-CoV-2 RNA or particles in water or human saliva across two different LAMP primer sets. A) Colorimetric LAMP reactions of viral RNA or particles in water or saliva, amplified with NEB Gene N-A primers or Lamb *et al*. primers as indicated. Independent biological triplicates are shown for each experiment. Values indicate number of viral genome equivalents per reaction. B) Quantification and comparison of approximate Limits of Detection (viral genome equivalents per reaction) were calculated across water or saliva, and primer sets. Limit of Detection was recorded as the lowest value with a clear colorimetric change from magenta to yellow. Inconclusive or undetected values were recorded as Not Detected. Quantification and comparison of approximate Limits of Detection (viral genome equivalents per reaction) were compared by RNA type, water or saliva, and primer sets. p-values indicate one-sided *t*-tests (saliva greater than water). Inconclusive or non-detected values were excluded. C) Limits of Detection in water were not significantly different across viral source (RNA or particles), or primer sets.

To neutralize or otherwise reduce inhibitors in human saliva, we tested several approaches that have been demonstrated to improve viral RNA detection in crude samples including saliva^13-17^. We tested simple dilution of particle-containing saliva into water, and various heat and chemical treatments. First, we found that dilution of saliva into water enabled sensitive detection of SARS-CoV-2 particles using LAMP (Figure 2A, top). A heat treatment of 55 °C for 15 minutes followed by 95 °C for 5 minutes further improved LAMP sensitivity (Fig. 2A, middle row). Identical heating steps plus the addition of proteinase K increased LAMP sensitivity relative to dilution alone (Fig. 2A, bottom row) but not markedly more than heat treatment. Importantly, we found that the combined heating steps above with and without proteinase K treatment improved SARS-CoV-2 particle detection in undiluted human saliva samples (Fig. 2B) and conferred a consistent limit of detection on the order of 10^2^ particles per reaction. This represents a 10,000-fold improvement in sensitivity over assays on untreated saliva. We experimented with additional heat and chemical pretreatments including the HUDSON protocol (heating unextracted diagnostic samples to obliterate nucleases)^14^ and various detergents, but all of these conditions decreased assay sensitivity or interfered with colorimetry (Supplementary Figure 2A-C). We also varied the amount of crude sample input to the LAMP reaction. We found that adding up to 8 μL of direct saliva was compatible with the assay but increased volume did not improve sensitivity (Sup. Fig. 2D).

**Figure 2:**
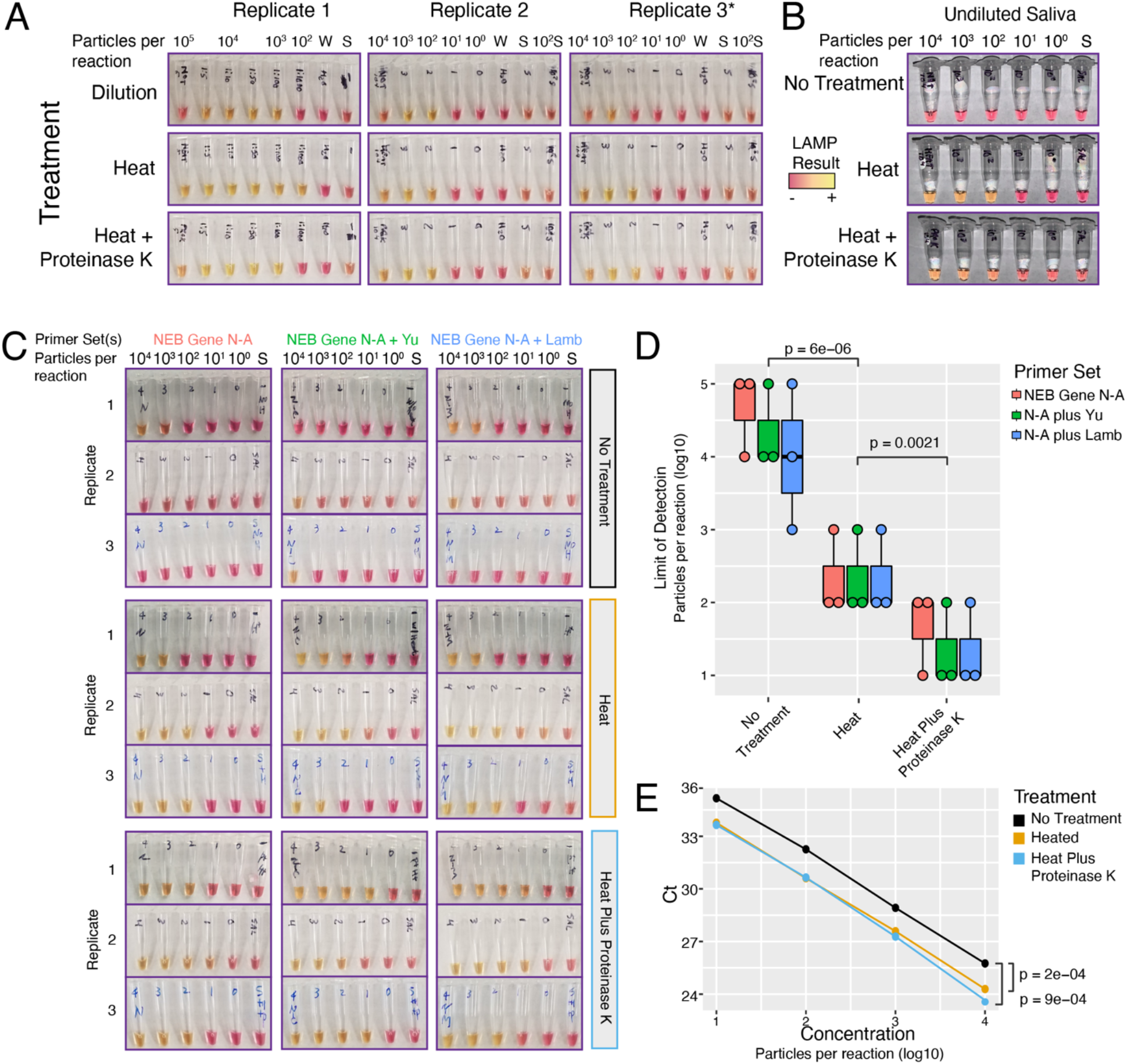
Dilution, heat, and Proteinase K treatments improve SARS-CoV-2 detection from saliva. A) Dilution of particle-containing saliva into water improved LAMP detection by at least two orders of magnitude from undetectable to ~10^3^ particles per reaction. Heat treatment and heat treatment plus proteinase K further increased LAMP sensitivity to ~10^2^ viral genome equivalents per reaction. *Replicate 3 used Lamb *et al*. primers but gave nearly identical results to NEB Gene N-A primers. B) Heat treatment with or without proteinase K increased LAMP sensitivity from 10^6^ to ~10^2^ viral genome equivalents in undiluted saliva. **C)** Multiplexed primers improved LAMP sensitivity. LAMP reactions using NEB Gene N-A primers alone or in combination with Yu *et al*. or Lamb *et al*. primers are shown. S = negative control saliva. Viral particles per reaction are indicated. D) Saliva pretreatments significantly improved LAMP sensitivity. Heat treatment improved limit of detection (*p* = 6e-6, *t*-test, two-tailed vs ‘No Treatment’). Proteinase K treatment further improved heat treatment (p = 0.002, *t*-test, two-tailed vs ‘Heat’). Multiplexed primers increased the frequency of detection at ~ 10^1^ particles / reaction. N = NEB Gene N-A. E) qRT-PCR on crude saliva using the CDC N1 probe showed increased sensitivity with either heat or proteinase K treatment (*p* < 1e-3 for either treatment, two-tailed paired *t*-test).

### Multiplexing LAMP Primer Sets

To further improve the accuracy of our assay, we sought to multiplex LAMP primer sets in a single reaction. Combining primers can potentially increase sensitivity through additive signals of simultaneous amplification reactions^18,19^. Including multiple primer sets will also confer diagnostic robustness against mutations that arise in the SARS-CoV-2 genome^20^. Non-specific primer interactions, however, could result in increased rates of false positives. We compared pairwise combinations of NEB Gene N-A primers with the other four primer sets targeting various regions across the SARS-CoV-2 genome. Encouragingly, all pairs of primer sets outperformed the NEB Gene N-A primer set alone, with no apparent increase in spurious background amplification (Supplementary Figure 3).

We next tested whether multiplexing primer sets could improve signal detection in untreated and heat and chemical treated particle-containing saliva (Figure 2C). As before, we found that heat treatment (55 °C for 15 minutes, 95 °C for 5 minutes) alone gave a marked improvement in SARS-CoV-2 particle detection from saliva (Fig. 2D, *p* < 1e-5, two-sided *t*-test). This effect was consistent across all tested primer sets. The same heat treatment plus proteinase K further improved assay sensitivity compared to heat alone (*p* < 0.003, two-sided *t*-test). Multiplexed primer sets slightly improved the sensitivity of the assay, increasing the frequency of detection in samples with ~10^1^ particles per reaction. At this sensitivity, the multiplexed LAMP assay would detect the vast majority of COVID-19 positive samples based on reported saliva viral loads (median ~10^2^-10^3^ viral copies per μL)^10,11^, and virtually all infectious individuals^21^. As viral loads peak around the time of symptom onset, LAMP would have the highest accuracy at this critical timepoint for isolating carriers^22^.

To determine whether our extraction-free protocol also improved the sensitivity of SARS-CoV-2 detection in a qRT-PCR-based assay, we performed qRT-PCR using the US Centers for Disease Control and Prevention (CDC) Gene N1 probe set directly on untreated and treated simulated saliva samples. We found that qRT-PCR had similar sensitivity to LAMP on crude samples, reliably detecting SARS-CoV-2 in all samples down to ~10^1^ particles per reaction (Fig. 2E). Reactions with 10° particles were not reliably detected in this assay. We observed strong improvements in cycle thresholds (Ct) using either heat alone or heat plus proteinase K (*p* < 1e-3, two-tailed paired *t*-tests). These experiments demonstrate that our saliva pretreatment increases the sensitivity of viral RNA detection by qRT-PCR by 3-4 fold. Taken together, our results show that a simple, extraction-free pretreatment protocol can significantly improve the limit of detection of downstream nucleic acid-based assays.

### Validation on Clinical Samples

We obtained saliva samples collected at day zero of hospital admission from five COVID-19 positive individuals. Samples were aliquoted into 3 tubes for either no treatment, heat inactivation (55 °C for 15 minutes, 95 °C for 5 minutes), or the same heat treatment plus proteinase K. Following pretreatment, the RT-LAMP reaction was performed with the NEB Gene N-A primers. Photographs were taken at 10 and 30 minutes to track colorimetric shifts over time (Figure 3A). An aliquot of 10^4^ viral particles per μL in saliva was used as a positive control. After 30 minutes, 4 of the five samples were clearly positive (yellow) in the heat plus proteinase K treated samples (Fig. 3A, bottom right). Positive and negative controls were positive and negative in all reactions at the 30-minute timepoint, allowing an interpretable readout of the assay. Untreated samples showed positivity for samples 3-5, and heat-alone pretreatment indicated positivity for samples 2-4. These differences may reflect altered ratios of free viral RNA to particle-associated RNA in the starting samples, which could be differentially affected by heat treatment. Heat releases RNA from viral particles and inactivates reaction inhibitors but can degrade free RNA. Proteinase K treatment may protect free RNA by inactivating nucleases.

**Figure 3:**
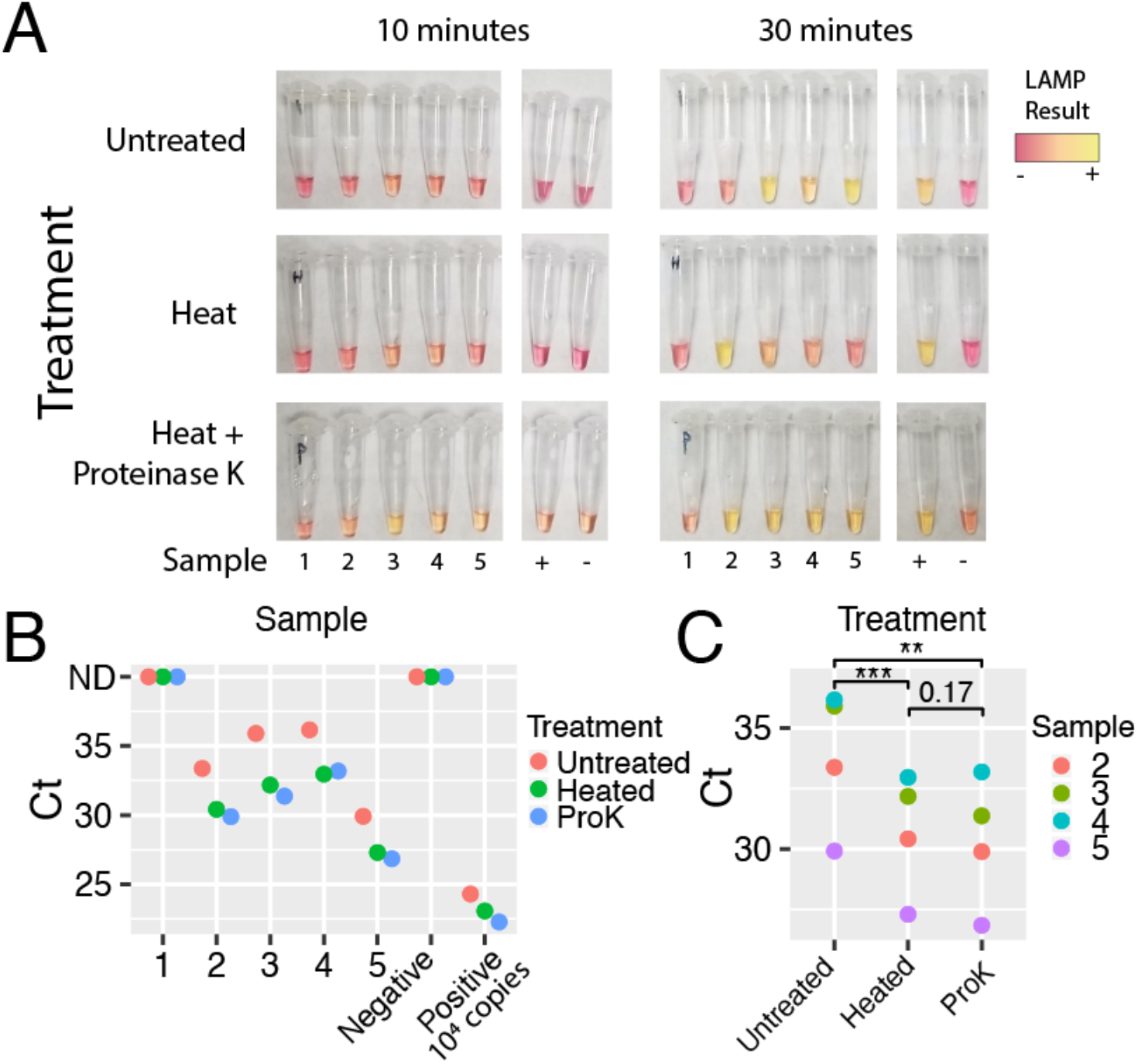
Preliminary validation of RT-LAMP and qRT-PCR on clinical saliva samples. A) Saliva samples from five COVID-19 positive individuals were subjected to the indicated pretreatment protocol followed by RT-LAMP. Heat = 55 °C for 15 minutes, 95 °C for 5 minutes, with or without proteinase K. Photographs for timepoints 10- and 30-minutes are shown. By 30 minutes, all positive controls were positive (yellow), and negative controls remained negative (magenta). 4 out of 5 samples pretreated with heat plus Proteinase K were called positive by colorimetric LAMP. B) qRT-PCR performed directly on crude pretreated saliva samples was qualitatively concordant with the LAMP assay. Samples 2-5 were positive (Ct < 40). Sample 1 and negative control gave no signal. C) Pretreatment protocols enhanced sensitivity of qRT-PCR (two-sided paired *t*-test, ** = *p* < 0.01, *** *p* < 0.001). ProK, proteinase K. Ct, cycle threshold. ND, not detected (Ct > 45).

Lacking corresponding gold-standard qRT-PCR values from these samples, we instead performed qRT-PCR with the CDC N1 probe directly on the untreated and treated samples, along with positive and negative controls. qRT-PCR results were qualitatively concordant with the RT-LAMP results, identifying the same 4 positives (Samples 2-5, Ct < 40, Fig. 3B). Sample 1 tested negative in all LAMP assays and qRT-PCR, and an attempt to purify RNA from this saliva sample was unsuccessful, suggesting the RNA in this sample was degraded prior to LAMP and qRT-PCR. Further work is needed to establish best practices for upstream handling of saliva specimens. Toward this end, we found that the LAMP reaction is compatible with samples diluted in phosphate buffered saline (PBS), TE buffer (10 mM Tris, 0.1 mM EDTA), diluted HUDSON buffer (2.5 mM TCEP, 0.5 mM EDTA), and the additive RNAsecure, a non-enzymatic ribonuclease inhibitor that does not require heat inactivation. (Supplementary Figure 4A).

As with LAMP, our pretreatment regimens significantly improved nucleic acid detection by qRT-PCR (Fig. 3C). Heat treatment (55 °C for 15 minutes, 95 °C for 5 minutes) with and without proteinase K improved viral detection compared to untreated samples (*p* < 0.01 and *p* < 0.001 respectively). Proteinase K did not further significantly improve sensitivity compared to heat alone. Extrapolation of viral loads in clinical saliva samples by comparing to quantitative positive controls yielded estimates of 1.4 x 10^1^ – 9.8 x 10^2^ particles per μL and demonstrates the sensitivity of our assay on real clinical samples.

### Establishing a High-throughput Quantitative Assay

To enable significant scale-up of testing capacity using the LAMP assay on saliva, we adapted our protocol to a 96-well plate format. Spectrophotometric plate scanning before and after the assay provided an unbiased, quantitative interpretation. Initial plate scanning was implemented to normalize for baseline colorimetric differences induced by variation in pH across saliva samples. Heat treatment (55 °C for 15 minutes, 95 °C for 5 minutes) with and without proteinase K enabled unbiased and sensitive detection of viral particles in saliva samples down to 10^2^ particles per reaction, with some detection at 10^1^ particles per reaction (Figure 4A).We next sought to establish a quantitative real-time fluorescent-based LAMP assay (qLAMP) using the DNA intercalating dye SYTO 9^23^. qLAMP offers several potential advantages over colorimetric LAMP including real-time reaction monitoring and melt curve analysis to discriminate false positives. We first benchmarked qLAMP using contrived samples of known amounts of viral particles in diluted saliva, and we determined that a cycle threshold of 50 (25 minutes) could reliably discriminate positive reactions from nonspecific amplifications (Figure 4B). We also found that the dye used in colorimetric LAMP can be directly added to the qLAMP master mix, so that qLAMP reactions set up in this fashion can be read out by colorimetry or fluorimetry (Figure 4C), and these produce qualitatively concordant results (Figure 4D). Spectrophotometry and real-time LAMP therefore represent two alternative modalities for high-throughput, unbiased LAMP implementation.

**Figure 4:**
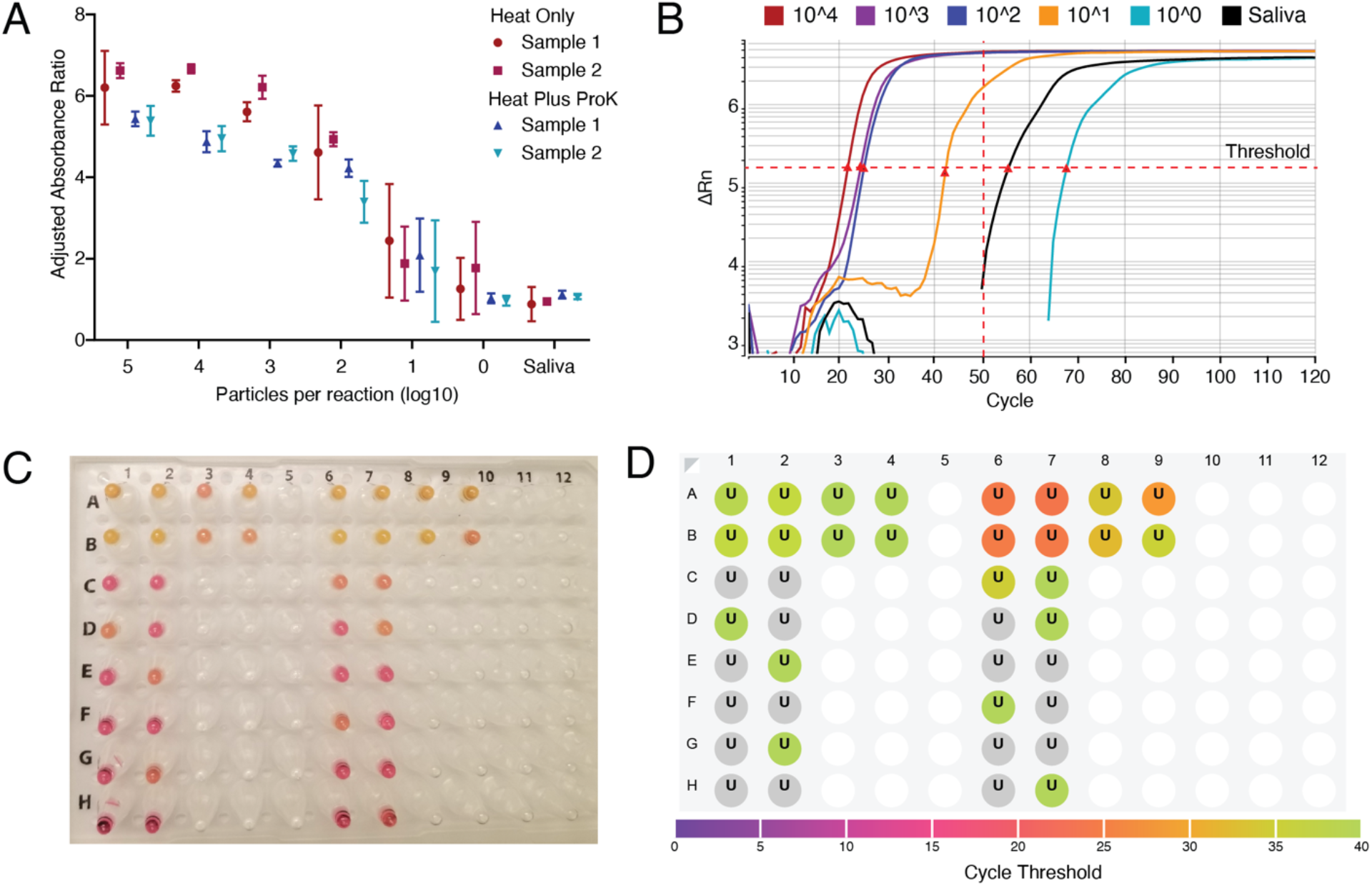
Establishing high-throughput LAMP assays with quantitative readouts. A) RT-LAMP assay was adapted to a high-throughput 96-well plate format with a quantitative absorbance readout, achieving a limit of detection < 10^2^ particles per reaction from saliva samples. Absorbance for 430 nM (yellow) and 560 nM (red) wavelengths was measured before and after the LAMP reaction and normalized to negative controls. Heat indicates 55 °C for 15 minutes, 95 °C for 3 minutes, with or without proteinase K (ProK). Two biological replicates were each run in triplicate. B) Real-time quantitative fluorescent LAMP results are shown for a dilution series of particles in saliva. Change in fluorescence (delta Rn) is monitored over 120 ‘cycles’ of 30-second incubations at 65 °C. Cycle thresholds (Cts), indicated by red triangles, represent the time at which total fluorescence reaches a given level. Samples with higher viral loads reach this threshold earlier. Nonspecific amplification may arise after 50 cycles, corresponding to 25 minutes. C) Colorimetric and D) fluorescent results for the same reaction show that the fluorescent dye does not interfere with colorimetric interpretation. Results are concordant with colorimetric LAMP, and fluorescent results are more quantitative.

### Improving Compatibility with Point-of-Care Testing

Isothermal LAMP is well-suited to point-of-care testing because it requires only a heat-source. We sought to make the saliva pretreatment compatible with a single isothermal heat source to reduce equipment requirements and facilitate point-of-care testing. Whereas a mild heat treatment consisting of 50 °C for 5 minutes and 64 °C for 5 minutes did not enable sensitive detection of SARS-CoV-2 in undiluted saliva (Sup. Fig. 2B), a 65 °C treatment for 15 minutes of saliva that had been diluted 1:1 in either TE (10 mM Tris, 0.1 mM EDTA) or modified HUDSON buffer (2.5 mM TCEP, 0.5 mM EDTA) improved detection (Sup. Fig. 4B). Addition of RNAsecure boosted assay sensitivity to 10^2^ viral particles per reaction (Sup. Fig 4B). SARS-CoV-2 in samples treated with a thermolabile version of proteinase K (inactivated by incubation at 65 °C for 10 minutes) was not detected in a sensitive fashion (Sup. Fig. 4C). We found that guanidine hydrochloride (40 mM) was compatible with the LAMP reaction and that both RNAsecure and primer multiplexing provided further enhancements to sensitivity (Sup. Fig. 4D). Pulse spinning pretreated saliva samples in a microfuge immediately prior to adding them to the LAMP reaction significantly improved assay reliability (Sup. Fig. 4E). While not quite as sensitive as pretreatments including a 95 °C heat step, these pretreatment methods enable the testing of saliva in colorimetric LAMP reactions using a single heat source, simplifying point-of-care testing.

### Re-optimizing LAMP assay

Given the improvements we observed in assay sensitivity using RNAsecure, guanidine, and sample dilution into TE, we sought to incorporate these modifications into our highest sensitivity protocol (55 °C for 15 minutes, 95 °C for 5 minutes). We increased the 55 °C stage to 65 °C for better inactivation of virus, RNases, and reaction inhibitors. During optimization experiments, we observed a low rate of non-specific amplification arising in experiments with multiplexed primers (Supplementary Figure 5A). We found that halving the primer concentrations or addition of betaine reduced this occurrence, but these diminished assay sensitivity (Sup. Fig. 5A-B). Computational re-analysis of LAMP primer sets indicated potential primer-primer interactions that could generate spurious products (Sup. Fig. 5C). To avoid such primer interactions, we switched to primer sequences redesigned by NEB targeting the nucleocapsid and envelope small membrane protein (E) genes (NEB-N2 and NEB-E1 primers)^24^.

We validated the performance of the new primer sets with both colorimetric LAMP and qLAMP. These experiments indicated the new NEB-N2 primer set outperformed the previous primers in both sensitivity and time to threshold (Supplementary Figure 6A,B). The addition of RNAsecure improved sensitivity across all primer sets (Sup. Fig. 6C). Next, we tried multiplexing the new primer sets with the previous primers. Multiplexing NEB-N2 and NEB-E1 gave the most consistent results with no false positives in saliva-only controls (Sup. Fig 6D). Guanidine improved both the speed and sensitivity of the LAMP reactions (*p* < 0.001, Sup. Fig. 6D-E).^24^

To calculate the limit of detection for our optimized assay, we performed the assay on dilution series of viral particles spiked into multiple donor saliva samples. In parallel, we tested whether proteinase K inclusion was beneficial. Our optimized assay was highly sensitive across all donors, detecting 96.9% of samples with 25 particles/μL (75 particles per reaction) with 100% specificity. (Table 1, Supplementary Figure 7A).

**Table 1:** Limit of Detection Analysis. SARS-CoV-2 viral particles spiked into three healthy donor saliva samples in two-fold dilutions were pretreated then measured using qLAMP. Positive or negative results were called using a cycle threshold below or above 50 respectively (See Sup. Fig. 7A).

| Limit of Detection Analysis |  |  |  |
| --- | --- | --- | --- |
| Amount (Particles/μL) | Replicates | Called Positive | Sensitivity (Specificity) |
| 100 | 32 | 32 | 100% |
| 50 | 32 | 32 | 100% |
| 25 | 32 | 31 | 96.9% |
| 12.5 | 32 | 27 | 84.4% |
| 6.25 | 32 | 23 | 71.9% |
| 3.125 | 32 | 10 | 31.3% |
| 0 | 32 | 0 | 100.00% |

Proteinase K significantly improved SARS-CoV-2 detection in samples with the lowest viral amounts (p = 0.006, Sup. Fig. 7B). To optimize assay compatibility with clinical samples that had already been diluted in PBS, we substituted TE instead of water in the LAMP master mix (Sup. Fig. 7C-D). This modification offers buffering against basal pH differences in saliva without affecting assay sensitivity. Including these samples, we recalculated the limit of detection on samples pretreated with heat and proteinase K. By this analysis, our optimized assay including proteinase K in the pretreatment achieved 100% sensitivity (24/24 samples) at 12.5 particles/μL and 91.7% sensitivity at 6.25 particles/μL, with 100% specificity on freshly prepared simulated samples. With these finalized pretreatment conditions, we re-tested the optimum amount of saliva that should be added to the reaction. We found that increasing amounts of pretreated saliva impeded reaction times (Sup. Fig. 7E). Based on this we recommend adding 1-4 μL of pretreated saliva in a 20 μL LAMP reaction. Reaction volumes and saliva amounts can be scaled up proportionally to increase assay sensitivity but at higher costs per reaction.

Finally, our optimized saliva pretreatment protocol again markedly improved the detection of SARS-CoV-2 by qRT-PCR (Sup. Fig. 7F). Compared to untreated saliva, heat and proteinase K pretreatment improved viral detection by an average 3.4-fold (*p* < 3e-9, *t*-test). qRT-PCR on direct saliva without RNA extraction is thus another viable option to overcome bottlenecks limiting widespread testing.

### Clinical Validation of Optimized Assay

We tested our optimized colorimetric RT-LAMP protocol on 30 additional clinical samples (20 positive and 10 negative samples). Samples had been previously collected, diluted in PBS, and frozen. We thawed the samples on ice and added 1/10^th^ volume of a solution containing RNAsecure and proteinase K to each sample. We then heated samples at 65 °C for 15 minutes, 95 °C for 5 minutes, and cooled them to 4 °C. We performed colorimetric LAMP and qRT-PCR on treated samples.

By naked-eye interpretation of colorimetric LAMP, 17/20 positive samples were correctly called positive and 9/10 negative samples were called negative (Figure 5A). The false positive result was found to be negative when re-tested with an alternate set of LAMP primers, indicating possible carry-over contamination of LAMP amplicons^25^. This can be prevented by including uracil (dUTP) and a uracil glycosylase in the reactions^26^, as is commonly implemented in clinical qRT-PCR. qLAMP Cts had strong overall agreement with colorimetric results. qLAMP Cts for five samples were too high to distinguish between weak positive signal or non-specific amplification (Fig. 5B, yellow circles, red numbered samples). qRT-PCR correctly called 19/20 positive samples (Fig. 5B, purple squares), and 10/10 negative samples (Table 2). qRT-PCR Cts were well-correlated with LAMP Cts (Fig. 5C). One positive sample was undetected by either method, suggesting sample degradation or very low viral levels. Future assays should incorporate an endogenous human RNA target such as RNase P to improve interpretation.

**Figure 5:**
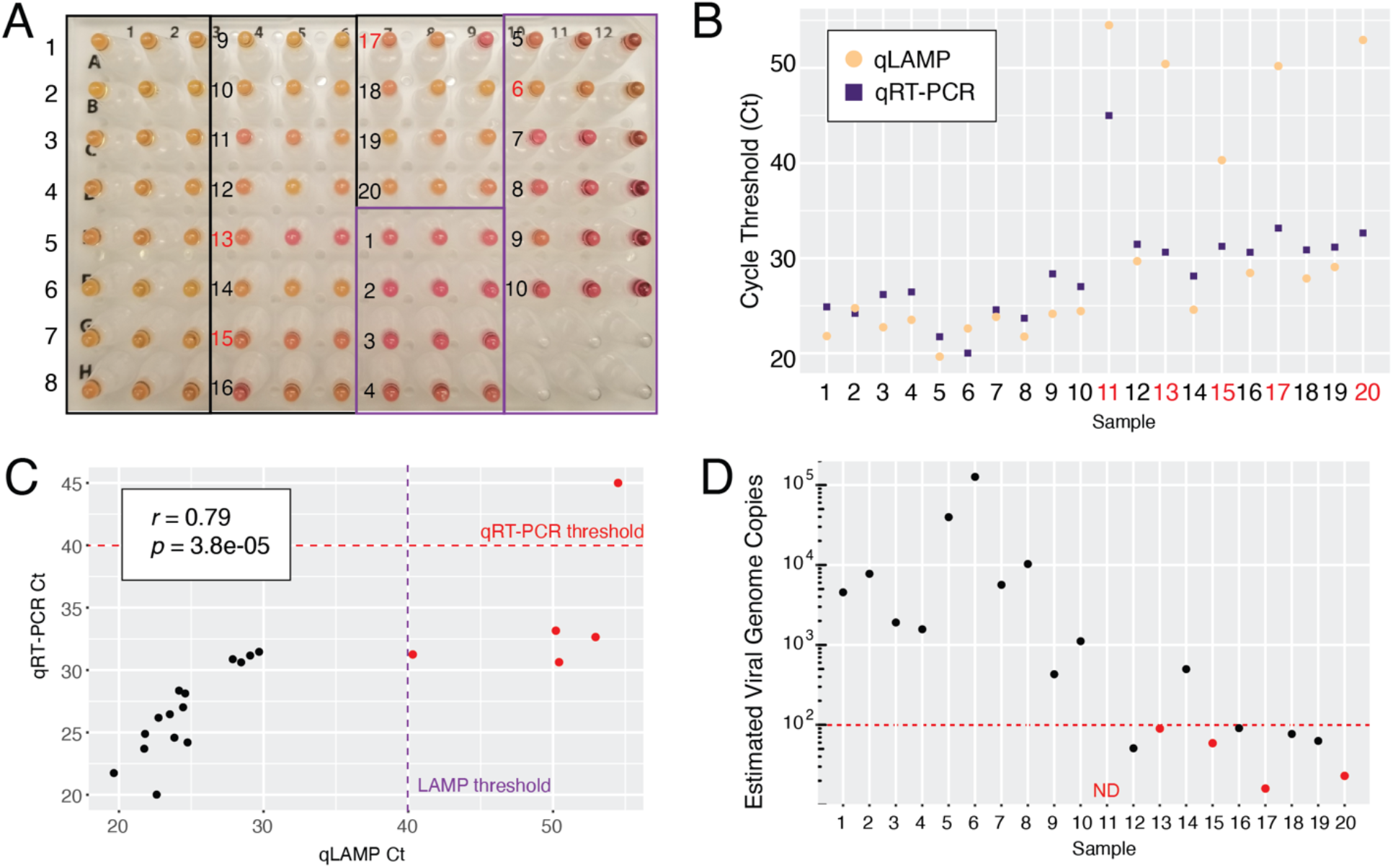
Clinical Validation of Optimized Colorimetric LAMP Assay and qRT-PCR. A) Colorimetric LAMP results on 30 clinical samples (20 positives, 10 negatives). Correct calls are numbered in black. Incorrect calls are numbered in red. B) qLAMP and qRT-PCR results on pretreated samples are shown for each sample. Red numbers indicate false negative calls by qLAMP. C) qLAMP and qRT-PCR Ct values were highly correlated. Thresholds of 40 cycles were used for determining positivity for LAMP and qRT-PCR as indicated. Samples negative by either of these methods are colored red. D) Estimates of viral genome copy number were made for each sample using a quantitative standard curve by qRT-PCR. Samples called negative by LAMP are colored red. Red-dotted line indicates 100 viral genomes. ND = Not Detected.

**Table 2:** Accuracy of LAMP and qRT-PCR assays on pretreated clinical saliva samples. * after re-testing.

|  | True Positive | False Negative | True Negative | False Positive | Sensitivity (%) | Specificity (%) | Accuracy (%) |
| --- | --- | --- | --- | --- | --- | --- | --- |
| **Colorimetric LAMP** | 17 | 3 | 10* | 0* | 85 (62.1-96.8) | 100 (69.1-100) | 90 (73.5-97.9) |
| **qRT-PCR** | 19 | 1 | 10 | 0 | 95 (75.1-99.9) | 100 (69.1-100) | 96.7 (82.8-99.9) |

We included a quantitative dilution series of particles in the qRT-PCR assay to estimate viral loads in clinical samples. The median viral copy number in positive samples was ~500 copies per μL (range 16-126,000). All of the false negatives in the LAMP assay had fewer than 100 estimated viral copies per μL which is likely below the threshold for infectiousness^21^. Nevertheless, among 8 samples below this level, LAMP achieved a 50% sensitivity. Above this level, the assay achieved 100% sensitivity. Therefore, colorimetric RT-LAMP on pretreated saliva samples without RNA extraction is a cheap, fast, and accurate method for SARS-CoV-2 testing.

## Discussion

Our proposed approach combines three promising avenues to enable rapid and widespread SARS-CoV-2 detection: 1) colorimetric RT-LAMP, 2) self-collected saliva specimens, and 3) compatibility with crude saliva samples without RNA extraction. This approach solves two major bottlenecks in massively scaling up COVID-19 nucleic acid testing: sample collection and RNA extraction, and it enables test result turnaround times less than an hour. Colorimetric RT-LAMP directly on saliva would facilitate rapid and frequent testing of pre- and asymptomatic carriers, enabling their isolation prior to unwitting viral transmission. Such testing is critical to curtailing the ongoing pandemic.

Due to its ease of use, rapid amplification of nucleic acids, high specificity arising from the use of six primers, and high tolerance of reaction inhibitors^8^, RT-LAMP has been widely used for pathogen detection. Sensitive diagnostic assays have been developed for viruses including Zika^19,27^ and such assays are being developed for SARS-CoV-2 by several groups including ours^3-7,18,28-32^. Speed, cost, turnaround time, and a simple colorimetric readout make RT-LAMP an effective solution to ramping up testing capacity. Further, because it does not require specialized equipment or training for performing or interpreting the assay, RT-LAMP is especially well-suited for point-of-care detection. Sample collection is currently limited by the reliance on NP swabs, which need to be carefully performed by a trained health-care worker and require the use of PPE. Mid-nasal swabs are a promising alternative to NP swabs because they can be self-administered, and contain high viral loads^33-35^. However, due to potential swab shortages, we instead focused on saliva due to its ease of collection and high viral load^11^.

Saliva is a challenging clinical matrix due to variability across individuals in pH and viscosity and the presence of reaction inhibitors^36^. Here, we have overcome these challenges and demonstrate a variety of saliva pretreatment protocols that enable sensitive detection of SARS-CoV-2 by both RT-LAMP and qRT-PCR without an RNA extraction step. Several groups are optimizing workarounds to avoid the RNA extraction step for nucleic acid-based SARS-CoV-2 testing while maintaining sensitivity^13,15,17,33^. Building on these efforts, we achieved a careful balance between the inactivation of reaction inhibitors and the preservation of viral RNA with heat and chemical pretreatment. A 1:1 dilution of saliva followed by treatment with RNAsecure and 65 °C incubation potently reduces reaction inhibitors, and can be implemented with a single heat source isothermal with the LAMP reaction in a point-of-care setting. The addition of proteinase K and a brief incubation at 95 °C further reduces inhibitors, eases saliva handling, and improves assay sensitivity. For centralized lab testing in a high-throughput format, this pretreatment protocol can be coupled with colorimetric LAMP with a spectrophotometric or fluorescent readout, or with qRT-PCR. In a set of 30 clinical saliva samples, both colorimetric LAMP and qRT-PCR on treated samples performed with high accuracy. For clinical assay implementation, samples should be tested in duplicate across multiple primer sets targeting the SARS-CoV-2 genome and an internal human RNA control. Uracil incorporation and the addition of uracil DNA glycosylase should be used to prevent carryover contamination and false-positive results.

Frequent screening of employees, especially health-care workers, is feasible with this approach due to its rapid turnaround, low complexity, and low cost. As viral loads are likely correlated with transmission, only weakly infectious carriers risk false negative results by this assay which is also true for the current gold standard. Colorimetric RT-LAMP on saliva has broad potential to increase COVID-19 screening speed and capacity and has high flexibility in implementation depending on equipment availability. Similar LAMP-based approaches by others have also been validated on clinical samples, deployed for disease surveillance, and adapted for potential home testing and point-of-care use^31,32,37^. Moreover, the FDA has now granted Emergency Use Authorization to several LAMP-based assays, validating its utility to expand testing capacity.

In summary, we have optimized saliva pretreatment to enable sensitive SARS-CoV-2 detection from unpurified saliva samples in an optimized colorimetric RT-LAMP reaction and qRT-PCR. This optimization overcomes the burden of RNA extraction, and alleviates some of the time and labor bottlenecks of the current gold standard nucleic acid-based tests. Because of the flexibility of implementation and read-out, our assay can be deployed as a point-of-care test or in a centralized laboratory facility. Our extensive optimizations have enabled reliable detection below ~10^2^ viral genomes per reaction from saliva samples. Using both colorimetric LAMP and qRT-PCR, we achieved high accuracy on clinical saliva specimens without RNA extraction.

## Methods

### LAMP Reactions

All LAMP reactions were performed following New England Biolab’s recommended protocol using WarmStart Colorimetric LAMP 2X Master Mix (NEB, Massachusetts USA, M1800L). 20 μL reactions containing 10 μL LAMP master mix, 2 μL of 10X primer mix (2 μM F3 and B3, 16 μM Forward Inner Primer (FIP) and Backward Inner Primer (BIP), and 4 μM of Loop Forward (LF) and Loop Backward (LB) primers (25 or 100 nmol scale, IDT), 5 μL nuclease-free water, and 3 μL samples. LAMP reactions were incubated at 65 °C using BioRad DNA Engine thermocyclers for 30-60 minutes. Photographs of samples laid on white sheets of paper were taken with cell phone cameras.

### SARS-CoV-2 Standards and Controls

*In vitro* transcribed RNA standards were prepared as described^12^. https://www.protocols.io/view/generation-of-sars-cov-2-rna-transcript-standards-bdv6i69e In brief, gBlocks (IDT) corresponding to SARS-CoV-2 regions targeted by LAMP primer sets were PCR amplified and *in vitro* transcribed using MEGAshortscript T7 Transcription Kit (ThermoFisher). RNA products were column-purified using Monarch RNA Cleanup kit (NEB) and quantified using Qubit. High concentration stock solutions were prepared, aliquoted for individual use, and frozen at -80 °C to prevent multiple freeze-thaws.

Heat inactivated SARS-CoV-2 particles were acquired from the CDC through BEI Resources. Viral stock concentrations were 1.16 x 10^6^ particles per μL. 10 μL particles were resuspended in 90 μL water or saliva to make 10^5^ particles/ μL stock. Alternatively, 10 μL particles were added into 990 μL saliva to make 10^4^ particles/ μL stocks which were individually aliquoted to reduce freeze-thaws.

DNA plasmid coronavirus controls corresponding to SARS-CoV-2 and MERS were obtained from IDT as plasmid DNA solutions. nCoV-N control: Severe acute respiratory syndrome coronavirus 2 isolate Wuhan-Hu-1, complete genome (GenBank: NC_045512.2). MERS control: Middle East respiratory syndrome-related coronavirus isolate KNIH/002_05_2015, complete genome (GenBank: MK796425.1)

### Saliva Pretreatments

HUDSON (heating unextracted diagnostic samples to obliterate nucleases)^14^ was performed as described using Tris(2-carboxyethyl)phosphine hydrochloride (TCEP, 100 mM) and EDTA (1 mM) final concentrations. A mild heat treatment consisting of 50 °C for 5 minutes and 64 °C for 5 minutes was employed. Neutral pH TCEP was also investigated, but still caused an instant colorimetric shift in LAMP reactions. While diluted TCEP/EDTA was compatible with LAMP, its ability to inactivate nucleases or lyse viral particles remains to be validated.

In early experiments, Proteinase K from NEB (#P8107S) was added to saliva at 1/10 volume (5 μL in 50 μL saliva). Later, this was used at 100X in samples diluted 1:1 in TE buffer with 1X RNAsecure (25X, ThermoFisher, AM7006). We performed heat inactivation of Proteinase K for 5 minutes at 95 °C. We also tested thermolabile proteinase K from NEB (P8111S). Samples were incubated at 37 °C for 10 minutes followed by proteinase K inactivation at 65 °C for 10 minutes.

### qRT-PCR

qRT-PCR reactions were performed according to CDC Emergency Use Authorization guidelines using TaqPath 1-Step RT-qPCR Master Mix GC (ThermoFisher, A15300) and the nCoV-N1 probe from the 2019-nCoV RUO Kit (IDT). Reactions were performed on Quantstudio 3 and 6 Real-Time PCR systems (ThermoFisher) and analyzed using the Quantstudio Design & Analysis Software (v2.3.3, ThermoFisher). When using crude saliva as input, 3 μL was used as input to match the LAMP protocol.

### High-Throughput Colorimetric Assay

Assay scale-up was performed in a 96-well plate format (BioRad 96-well skirted PCR plate) with only minor modifications to the LAMP reaction. 4 μL of saliva samples were used in 25 μL total volume reactions. Heat treatment (55 °C for 15 minutes, 95 °C for 3 minutes) and proteinase K treatment were identical to single tube format. RNasin (Promega) was included in these samples. Samples were run in technical triplicate at each dilution.

Utilizing a BioTek Epoch microplate spectrophotometer, sample plates are scanned prior to heating, designated as the pre-read, to establish “background” due to variations in color related to individual saliva samples. After heating at 65 °C for 30 minutes, the plate was read a second time, designated as the post-read. Each read takes approximately 1 minute. The reader software returns endpoint absorbance for 430 nM and 560 nM wavelengths that measure positive and negative reaction results, respectively. Analytically, the ratio of 430 nM to 560 nM is computed for the pre-and post-read scans. For each well, the pre-read ratio is subtracted from the post-read ratio to establish a background subtracted value. The mean and standard deviation of the background subtracted negative controls (saliva + PBS) were computed from 12 wells across the plate. For each replicate, an adjusted absorbance ratio is computed by taking the ratio of each sample to the average negative control value (background subtracted sample ratio / background subtracted negative control ratio).

### Quantitative real-time LAMP (qLAMP)

qLAMP was performed by adding the DNA binding dye SYTO 9 (ThermoFisher) at a concentration of 1 μM to the colorimetric LAMP reaction and running the reaction on a QuantStudio 3 or 6 Real-Time PCR system. Machines were programmed to run 90 or 120 isothermal cycles of 30 seconds at 65 °C. qLAMP Cts can be converted to minutes by dividing by 2. After isothermal LAMP cycles, reactions were slowly ramped up to 95 °C for inactivation and melt-curve analysis.

### Data Analysis

Most data were analyzed and plotted in R (v3.5.1)^38^ using ggplot2^39^. qLAMP experiments were analyzed using the Quantstudio Design & Analysis Software (v2.3.3, ThermoFisher), or exported for analysis in R.

### LAMP Primers

NEB_orf1a-A-F3   CTGCACCTCATGGTCATGTT NEB_orf1a-A-B3   AGCTCGTCGCCTAAGTCAA NEB_orf1a-A-FIP   GAGGGACAAGGACACCAAGTGTATGGTTGAGCTGGTAGCAGA NEB_orf1a-A-BIP   CCAGTGGCTTACCGCAAGGTTTTAGATCGGCGCCGTAAC NEB_orf1a-A-LF   CCGTACTGAATGCCTTCGAGT NEB orf1a-A-LB   TTCGTAAGAACGGTAATAAAGGAGC NEB_geneN-A-F3   TGGCTACTACCGAAGAGCT NEB_geneN-A-B3   TGCAGCATTGTTAGCAGGAT NEB_geneN-A-FIP   TCTGGCCCAGTTCCTAGGTAGTCCAGACGAATTCGTGGTGG NEB_geneN-A-BIP   AGACGGCATCATATGGGTTGCACGGGTGCCAATGTGATCT NEB_geneN-A-LF   GGACTGAGATCTTTCATTTTACCGT NEB_geneN-A-LB   ACTGAGGGAGCCTTGAATACA El-Tholoth_orf1ab-F3   TGCTTCAGTCAGCTGATG El-Tholoth_orf1ab-B3   TTAAATTGTCATCTTCGTCCTT El-Tholoth_orf1ab-FIP   TCAGTACTAGTGCCTGTGCCCACAATCGTTTTTAAACGGGT El-Tholoth_orf1ab-BIP   TCGTATACAGGGCTTTTGACATCTATCTTGGAAGCGACAACAA El-Tholoth_orf1ab-LF   CTGCACTTACACCGCAA El-Tholoth_orf1ab-LB   GTAGCTGGTTTTGCTAAATTCC Lamb_F3   TCCAGATGAGGATGAAGAAGA Lamb_B3   AGTCTGAACAACTGGTGTAAG Lamb_FIP   AGAGCAGCAGAAGTGGCACAGGTGATTGTGAAGAAGAAGAG Lamb_BIP   TCAACCTGAAGAAGAGCAAGAACTGATTGTCCTCACTGCC Lamb_LF   CTCATATTGAGTTGATGGCTCA Lamb LB   ACAAACTGTTGGTCAACAAGAC Yu orf1ab-F3   CCACTAGAGGAGCTACTGTA Yu_orf1ab-B3   TGACAAGCTACAACACGT Yu_orf1ab-FIP   AGGTGAGGGTTTTCTACATCACTATATTGGAACAAGCAAATTCTATGG Yu_orf1ab-BIP   ATGGGTTGGGATTATCCTAAATGTGTGCGAGCAAGAACAAGTG Yu_orf1ab-LF   CAGTTTTTAACATGTTGTGCCAACC Yu_orf1ab-LB   TAGAGCCATGCCTAACATGCT NEB_N2-F3   ACCAGGAACTAATCAGACAAG NEB_N2-B3   GACTTGATCTTTGAAATTTGGATCT NEB_N2-FIP   TTCCGAAGAACGCTGAAGCGGAACTGATTACAAACATTGGCC NEB_N2-BIP   CGCATTGGCATGGAAGTCACAATTTGATGGCACCTGTGTA NEB_N2-LF   GGGGGCAAATTGTGCAATTTG NEB_N2-LB   CTTCGGGAACGTGGTTGACC NEB_E1-F3   TGAGTACGAACTTATGTACTCAT NEB_E1-B3   TTCAGATTTTTAACACGAGAGT NEB_E1-FIP   ACCACGAAAGCAAGAAAAAGAAGTTCGTTTCGGAAGAGACAG NEB_E1-BIP   TTGCTAGTTACACTAGCCATCCTTAGGTTTTACAAGACTCACGT NEB_E1-LB   GCGCTTCGATTGTGTGCGT NEB_E1-LF   CGCTATTAACTATTAACG

### Clinical Samples

Saliva samples were collected at day zero of hospital admission from six COVID-19 positive individuals. Saliva samples were diluted 1:1 in phosphate buffered saline to facilitate pipetting and then frozen. Samples were then thawed and heat-treated at 56 °C for 30 minutes to inactivate the majority of live virus^40^. Samples were then re-frozen. Samples were thawed on ice prior to LAMP and qRT-PCR reactions.

Additional samples were collected similarly and diluted 1:1 in PBS. These samples were not heat inactivated prior to our assay but underwent several rounds of freeze-thaws.

### Ethics Approval

Saliva collection was approved by the institutional review board at Washington University School of Medicine (WU350, IRB#202003085). Informed consent was obtained for all participant samples.

## Data Availability

All relevant data are contained within the paper.

## Author Contributions

MAL, XC, SJL, CSS, LCB, MNW, and WJB developed and optimized the LAMP assay with significant intellectual contribution from MH, RDM, RSF, RDH, JM. Clinical samples were obtained, prepared, and tested by MAL, CCF, SJL, and RSF. Spectrophotometric assay scale-up and quantitative read-out were implemented and analyzed by CSS, LCB, MH, and RDH. Fluorescent assay scale-up was implemented and analyzed by MAL, XC, SJL, MNW, and WJB. MAL analyzed the data and prepared the manuscript, with editing and revision by all authors.

## Acknowledgments

The following reagent was deposited by the Centers for Disease Control and Prevention and obtained through BEI Resources, NIAID, NIH: SARS-Related Coronavirus 2, Isolate USA-WA1/2020, Heat Inactivated, NR-52286.

These studies were supported by donations to the WUSM COVID Research fund, the Department of Genetics and the McDonnell Genome Institute, and NIH grants P30 CA91842 and UL1 TR000448. We thank the Alvin J. Siteman Cancer Center at Washington University School of Medicine (WUSM) and Barnes-Jewish Hospital, the Institute of Clinical and Translational Sciences (ICTS), and the Tissue Procurement Core, which provided saliva samples. The Siteman Cancer Center is supported in part by an NCI Cancer Center Support Grant #P30 CA091842 and the ICTS is funded by NCATS Clinical and Translational Science Award (CTSA) program grant #UL1 TR002345.

## Supplementary Figures

**Supplementary Figure 1:**
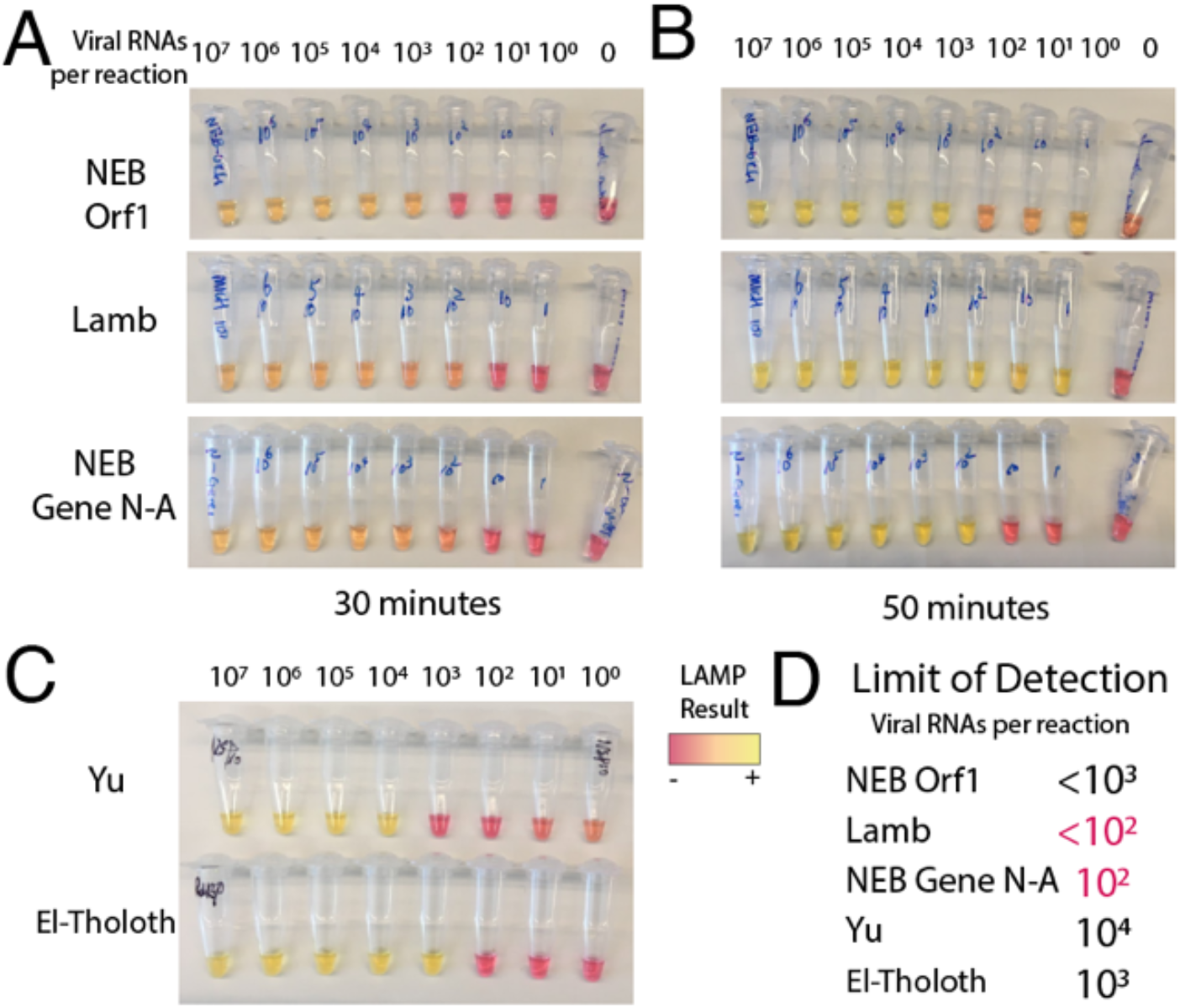
Testing LAMP primer sets targeting five separate regions of the SARS-CoV-2 genome. A) LAMP reactions produced colorimetric read-outs after 30 minutes for primer sets 1-3. Values indicate viral genome equivalent number of RNA per reaction. B) After 1 hour, some of the reactions became harder to interpret, as negative controls started turning yellow. C) Colorimetric LAMP assay for primer sets 4 and 5 after 1 hour. D) Approximate limits of detection (sensitivity to detect N number of viral genomes per reaction) were recorded, with primer sets 2 and 3 displaying the highest sensitivity, and no background amplification.

**Supplementary Figure 2:**
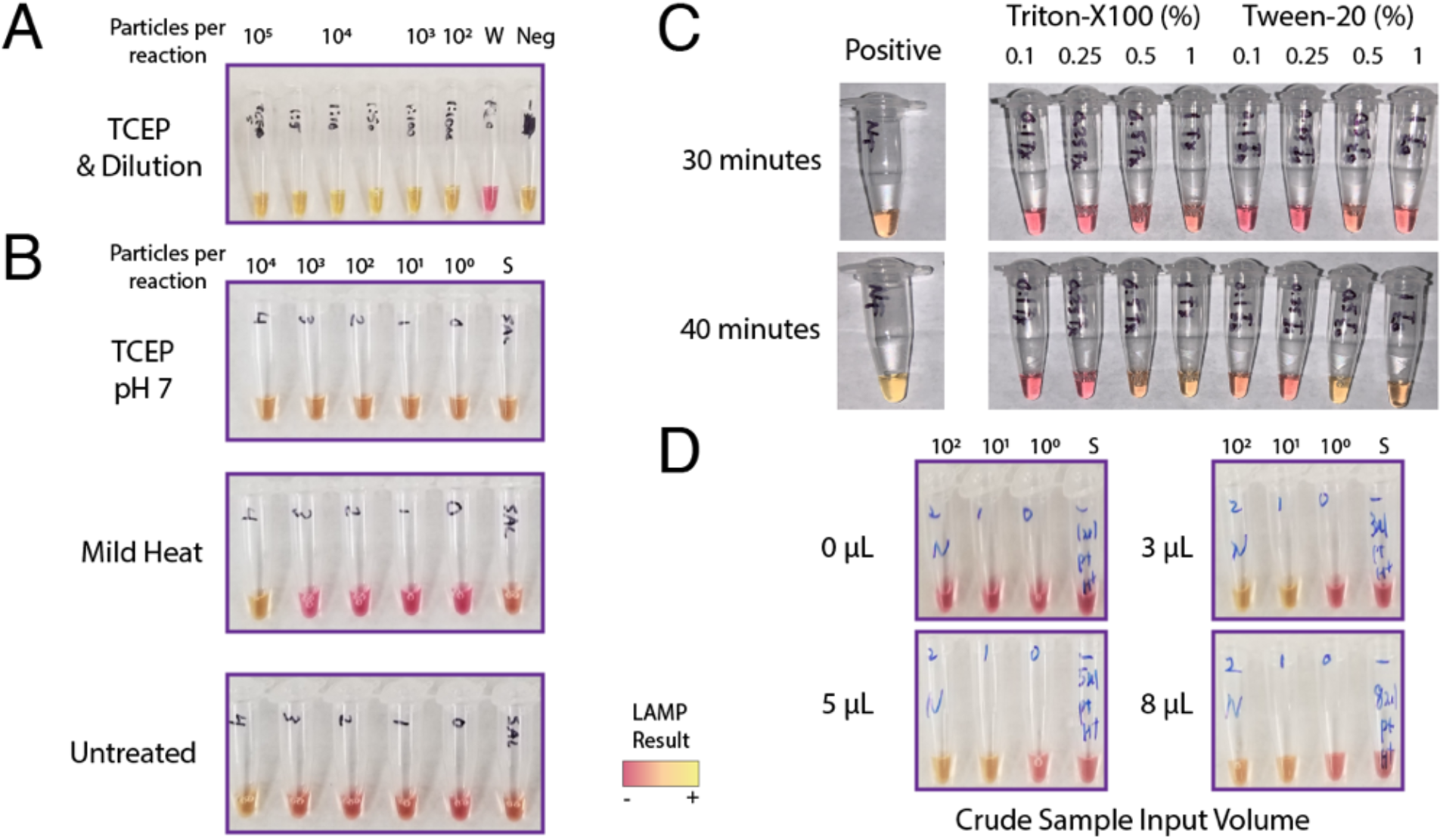
Evaluating heat and chemical pretreatments and reaction volume to increase detection of viral particles from saliva. A) The HUDSON method saliva heat and chemical method was applied but TCEP pH caused false colorimetric shift. B) Neutral pH formulated TCEP still induced colorimetric shift without viral RNA. Mild heat was not sufficient to improve particle detection, as it matched untreated conditions. C) Low levels of detergents Triton-X and Tween-20 inhibited LAMP detection at 30 minutes. D) The amount of crude saliva input to the LAMP reaction was varied. 3 uL was optimal to differentiate between positive and negative samples at 30 minutes. Increased volume did not increase the speed or sensitivity of the reactions. W = water. Neg = negative control saliva.

**Supplementary Figure 3:**
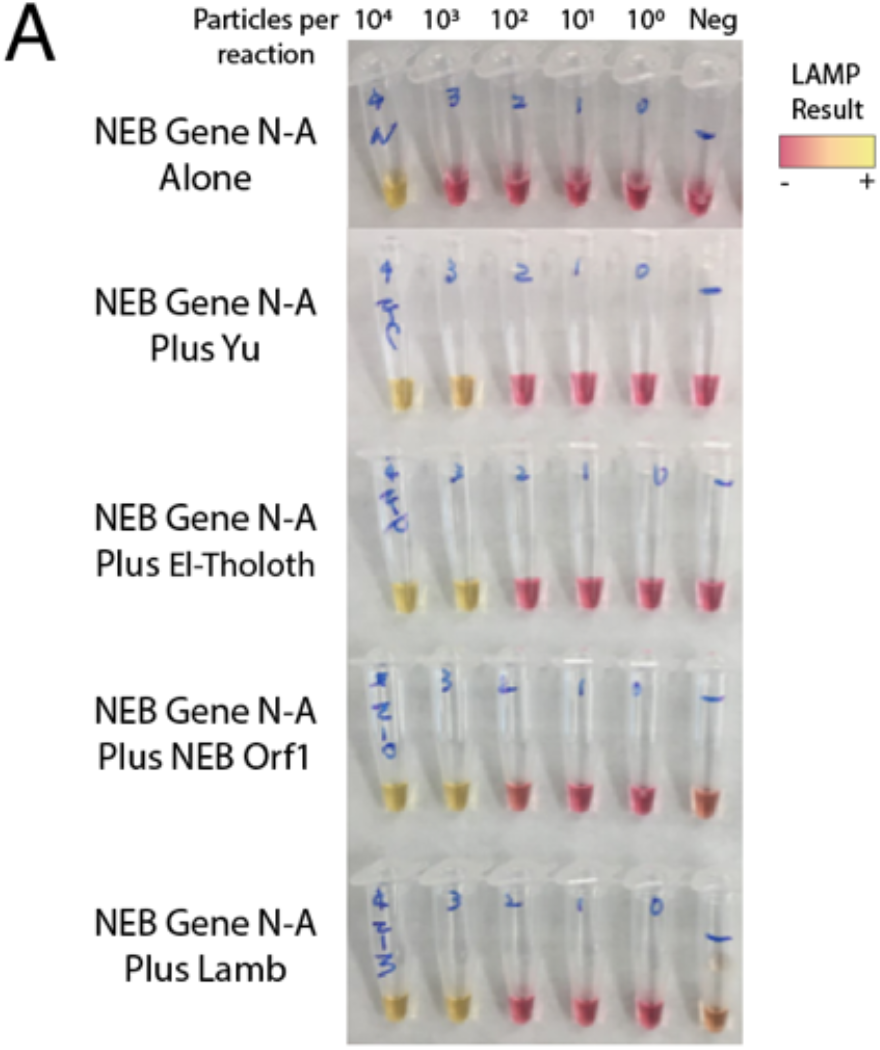
Multiplexing LAMP primer sets improves sensitivity by up to an order of magnitude. A) Colorimetric LAMP reactions using pairs of primer sets all increase detection sensitivity versus the single NEB Gene N-A primer set alone, with no increase in spurious amplification at 30 minutes. Neg = negative control saliva.

**Supplementary Figure 4:**
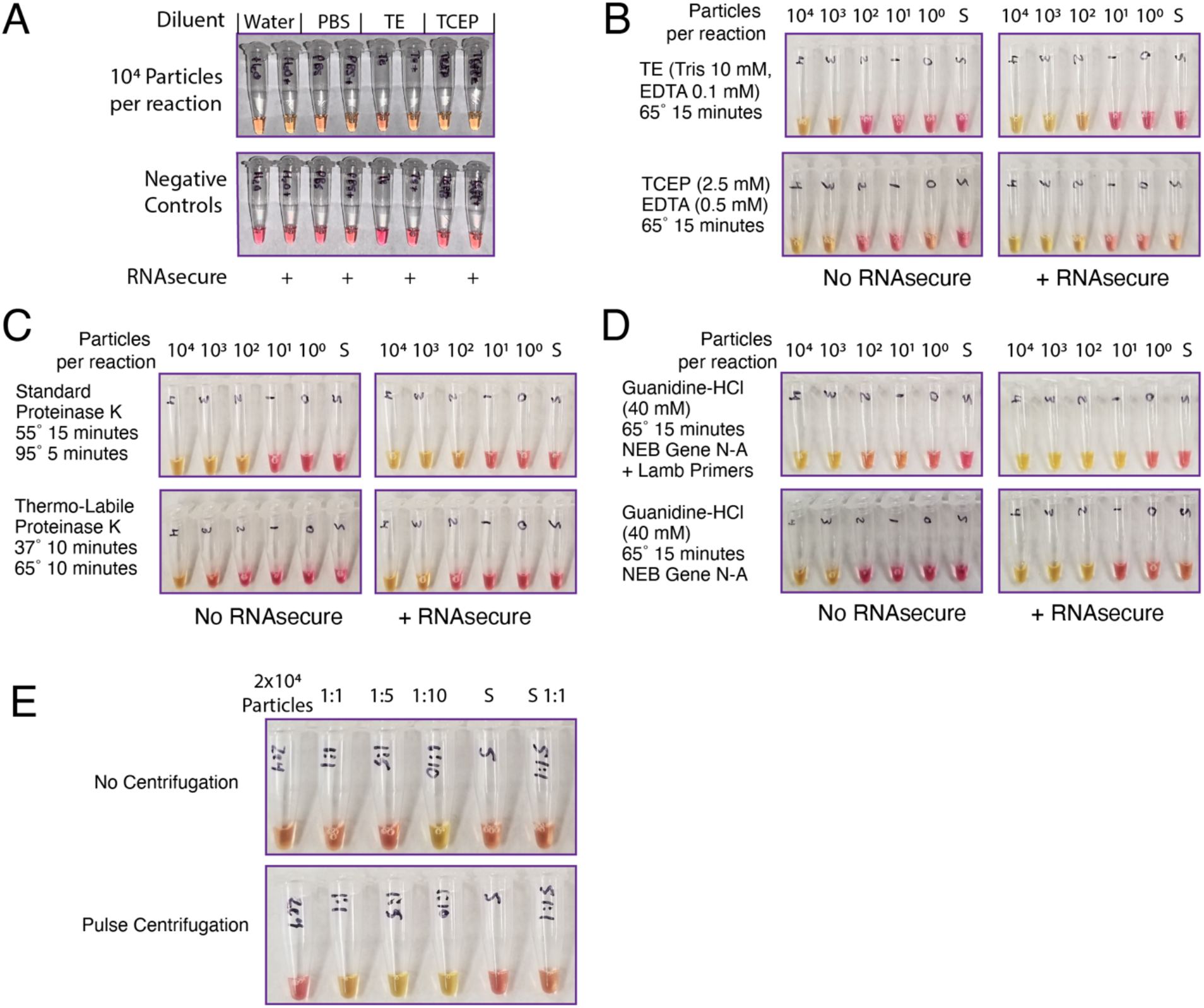
Establishing isothermal pretreatment methods to improve compatibility with point-of-care testing. A) Stabilizing buffers and ribonuclease inhibitor RNAsecure are compatible with LAMP. Colorimetric LAMP reactions proceed with saliva diluted 1:1 in either water, phosphate-buffered saline (PBS), TE (Tris 10 mM, EDTA 0.1 mM), and diluted TCEP (2.5 mM TCEP, 0.5 mM EDTA). B) A 15-minute isothermal (65 °C) heat pretreatment enables detection of 10^3^ viral particles from saliva. Addition of RNAsecure improves sensitivity down to 10^2^ viral particles. No additional benefit of diluted HUDSON reagents was seen compared to TE. C) Thermo-labile proteinase K treatment results in low sensitivity. While improved by RNAsecure, it performs worse than standard proteinase K with 95 °C inactivation, or mild heat treatments in (B). S = saliva control. D) Guanidium hydrochloride (40 mM) is compatible with the LAMP reaction. RNAsecure boosts the sensitivity in these conditions. Primer multiplexing in this buffer may further boost sensitivity to 10^1^ particles per reaction. All reactions performed with NEB Gene N-A primers, except in (D) which also included Lamb *et al*. primers as indicated. E) Pulse centrifugation in a microfuge after pretreatment improves the reliability of LAMP detection. S, saliva.

**Supplementary Figure 5:**
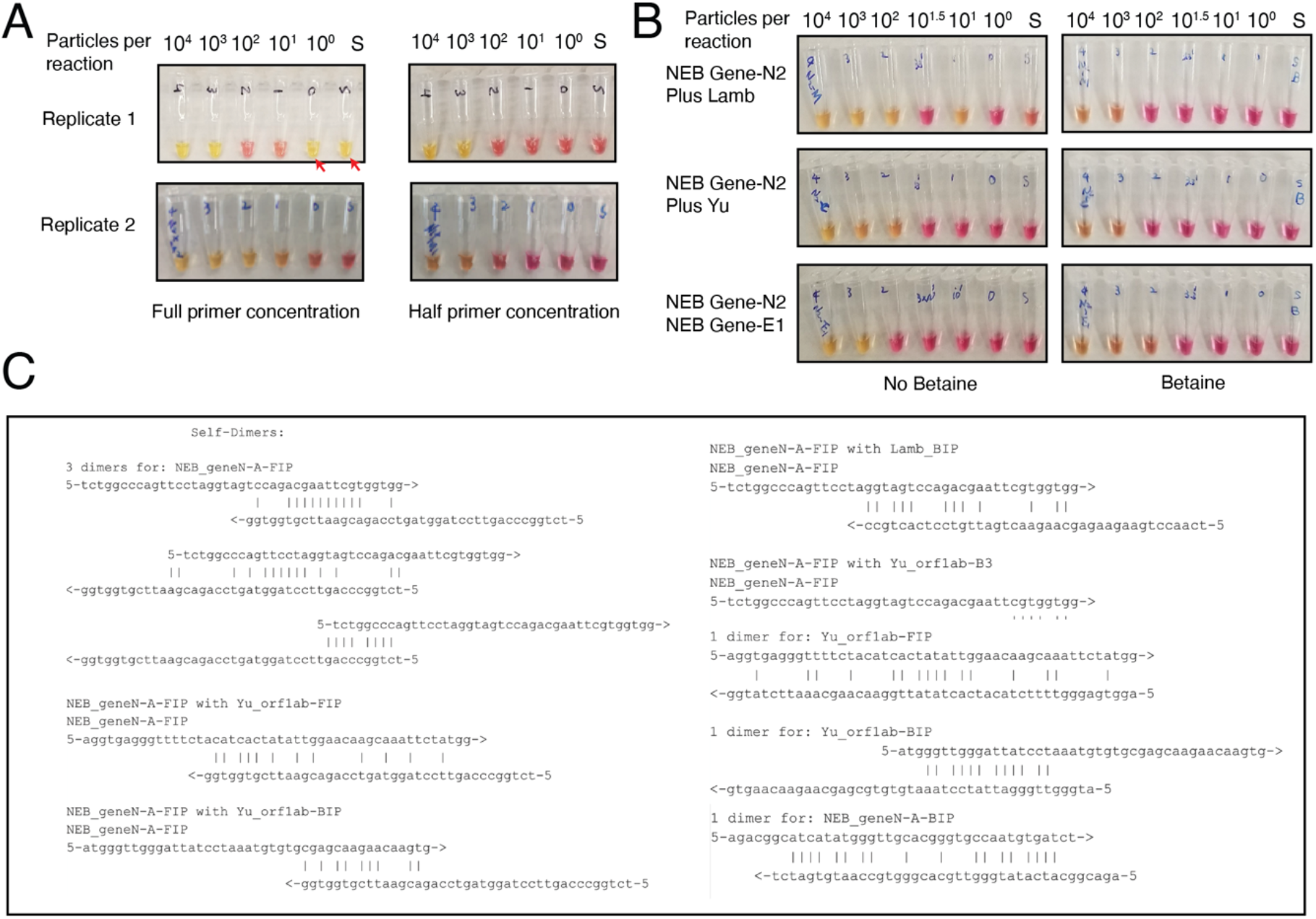
Non-specific amplification with multiplexed primer sets. A) Nonspecific amplicons were generated at low rates when using multiplexed primers. Red arrows indicate false positives. Halving the primer concentration reduced this occurrence, but reduced assay sensitivity. B) Betaine addition also reduced nonspecific amplification, but reduced assay sensitivity in all tested primer sets at 65. C) Computational analysis revealed predictions of potential primer-primer interactions across multiplexed primer sets.

**Supplementary Figure 6:**
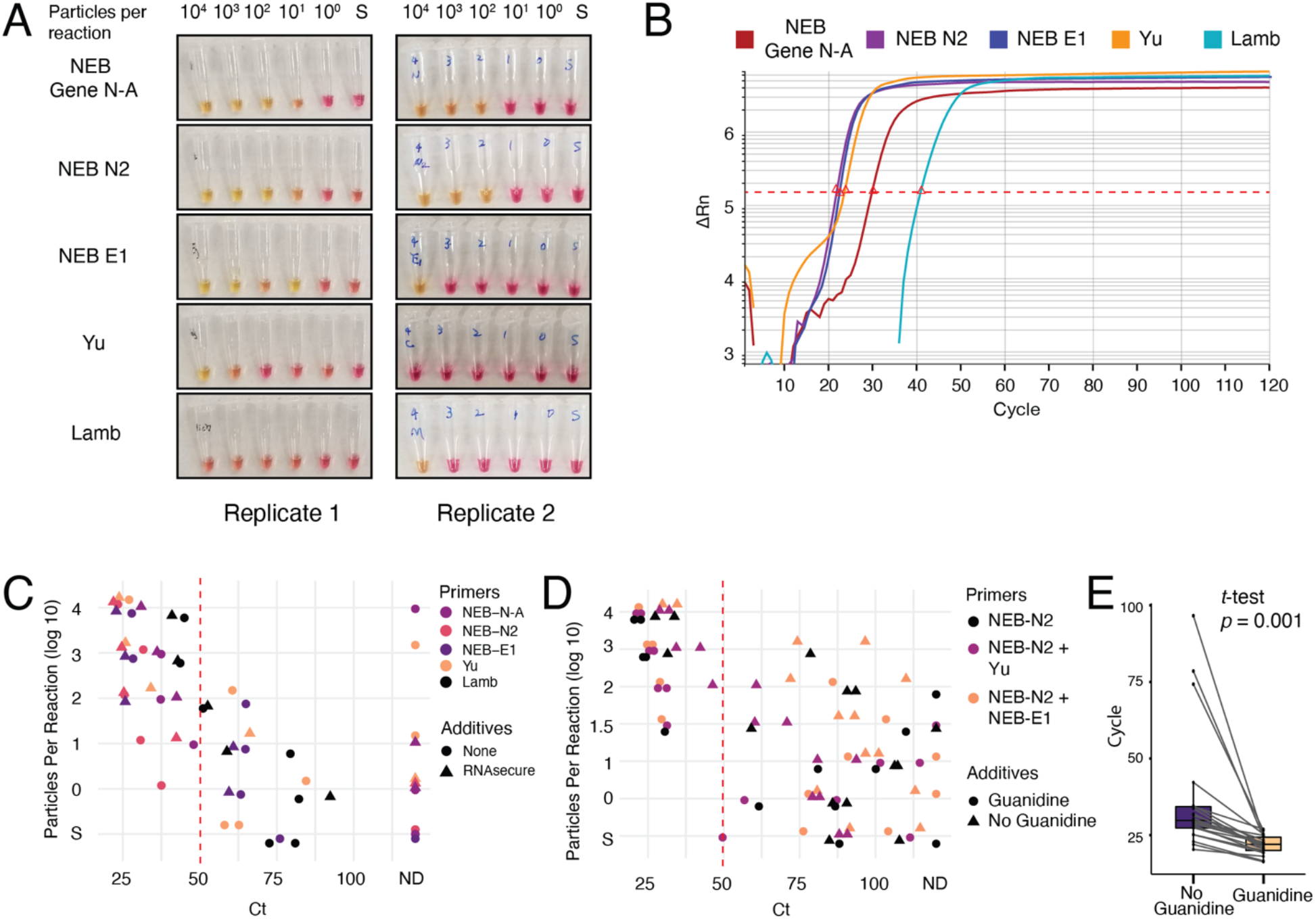
Validation and optimization of new primer set with colorimetric and fluorescent LAMP. A) Colorimetric LAMP reactions show that NEB N2 primers have high sensitivity. B) NEB N2 and NEB E1 primer sets had the fastest time to positive Cycle thresholds (Ct). Cts indicated with red triangle. Each cycle is 30 seconds. C) Plotting Ct vs particle concentration across primer sets and reaction additives reveals NEB-N2 primers are the most sensitive, and that RNAsecure generally improves LAMP reactions. Samples with Ct < 50 (dotted red line) are true positives. Non-specific amplification occurs after this time and is worse across different primer sets. ND, non-detected. D) Multiplexed NEB-N2 and NEB-E1 primer sets have high sensitivity and low background. Guanidine improves sensitivity. E) Guanidine improves time to positive result. Paired boxplot shows that guanidine lowers the Ct for almost all samples (*t*-test, *p* = 0.001).

**Supplementary Figure 7:**
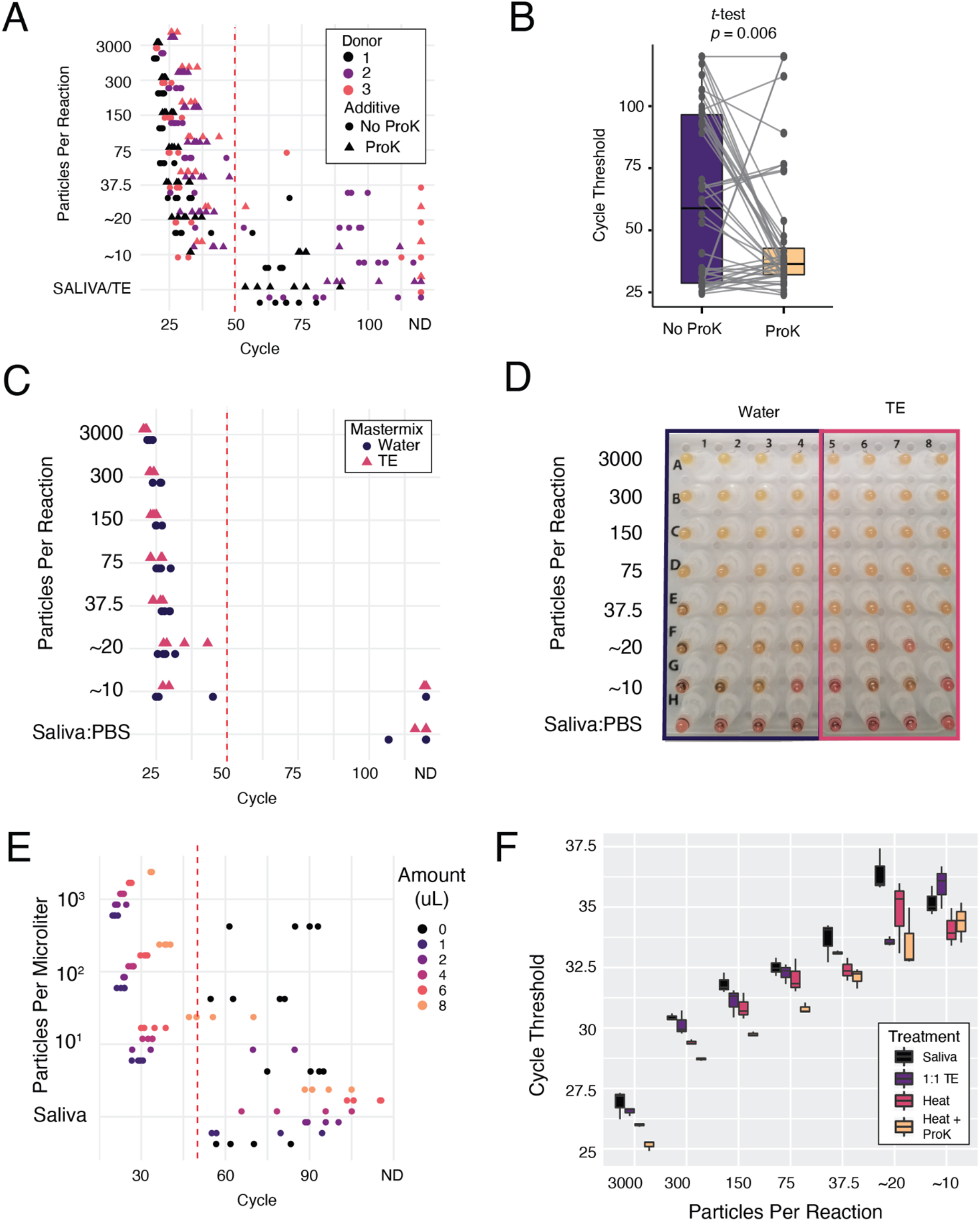
Final assay optimization, limit of detection analysis, and qRT-PCR. A) Limit of detection analysis on optimized assay. qLAMP results from serial dilution of particles across three donor saliva samples, with or without proteinase K. ProK, proteinase K. Samples with Cts less than 50 (dotted red line) are called positive. Results are tallied in Table 1. B) Paired box-plot of samples with less than 37.5 particles with or without proteinase K (ProK) shows that ProK reduces the mean and variability of Cts in these samples. C) Substituting TE for water in the fluorescent LAMP mastermix is compatible with samples that have been diluted in PBS. D) TE instead of water in the mastermix does not interfere with colorimetry. E) Increasing amounts of pretreated saliva containing three levels of virus were added to qLAMP reactions. Increasing amounts of pretreated saliva impede the LAMP reaction time. F) qRT-PCR on untreated and treated samples shows that heat with proteinase K treatment improves viral detection over untreated samples across all concentrations of virus. 1:1 TE samples are diluted but not heated.

## Notes

### Competing Interest Statement

Washington University in St. Louis has filed patent applications regarding this technology on behalf of the inventors.

### Summary of Updates

Colorimetric RT-LAMP assay optimization Establishment of fluorescent LAMP readout Additional validation of qRT-PCR assay on samples without RNA extraction Limit of Detection Analysis Validation on clinical specimens Recommendations for assay implementation

## References Cited

1. Notomi, T. et al. Loop-mediated isothermal amplification of DNA. Nucleic Acids Res. 28, E63 (2000).

2. Tanner, N. A., Zhang, Y. & Evans, T. C. Visual detection of isothermal nucleic acid amplification using pH-sensitive dyes. BioTechniques 58, 59–68 (2015).

3. Zhang, Y. et al. Rapid Molecular Detection of SARS-CoV-2 (COVID-19) Virus RNA Using Colorimetric LAMP. *medRxiv* 2020.02.26.20028373 (2020) doi:10.1101/2020.02.26.20028373.

4. Lamb, L. E., Bartolone, S. N., Ward, E. & Chancellor, M. B. Rapid Detection of Novel Coronavirus (COVID-19) by Reverse Transcription-Loop-Mediated Isothermal Amplification. *medRxiv* 2020.02.19.20025155 (2020) doi:10.1101/2020.02.19.20025155.

5. El-Tholoth, M., Bau, H. H. & Song, J. A Single and Two-Stage, Closed-Tube, Molecular Test for the 2019 Novel Coronavirus (COVID-19) at Home, Clinic, and Points of Entry. (2020) doi:10.26434/chemrxiv.11860137.v1.

6. Yu, L. et al. Rapid detection of COVID-19 coronavirus using a reverse transcriptional loop-mediated isothermal amplification (RT-LAMP) diagnostic platform. Clin Chem doi:10.1093/clinchem/hvaa102.

7. Rabe, B. A. & Cepko, C. SARS-CoV-2 Detection Using an Isothermal Amplification Reaction and a Rapid, Inexpensive Protocol for Sample Inactivation and Purification. *medRxiv* 2020.04.23.20076877 (2020) doi:10.1101/2020.04.23.20076877.

8. Francois, P. et al. Robustness of a loop-mediated isothermal amplification reaction for diagnostic applications. FEMS Immunology & Medical Microbiology 62, 41–48 (2011).

9. To, K. K.-W. et al. Consistent Detection of 2019 Novel Coronavirus in Saliva. Clin Infect Dis doi:10.1093/cid/ciaa149.

10. To, K. K.-W. et al. Temporal profiles of viral load in posterior oropharyngeal saliva samples and serum antibody responses during infection by SARS-CoV-2: an observational cohort study. The Lancet Infectious Diseases 0, (2020).

11. Wyllie, A. L. et al. Saliva is more sensitive for SARS-CoV-2 detection in COVID-19 patients than nasopharyngeal swabs. *medRxiv* 2020.04.16.20067835 (2020) doi:10.1101/2020.04.16.20067835.

12. Vogels, C. B. F. et al. Analytical sensitivity and efficiency comparisons of SARS-COV-2 qRT-PCR assays. *medRxiv* 2020.03.30.20048108 (2020) doi:10.1101/2020.03.30.20048108.

13. Marzinotto, S. et al. A streamlined approach to rapidly detect SARS-CoV-2 infection, avoiding RNA extraction. *medRxiv* 2020.04.06.20054114 (2020) doi:10.1101/2020.04.06.20054114.

14. Myhrvold, C. et al. Field-deployable viral diagnostics using CRISPR-Cas13. Science 360, 444–448 (2018).

15. Bruce, E. A. et al. DIRECT RT-qPCR DETECTION OF SARS-CoV-2 RNA FROM PATIENT NASOPHARYNGEAL SWABS WITHOUT AN RNA EXTRACTION STEP. *bioRxiv* 2020.03.20.001008 (2020) doi:10.1101/2020.03.20.001008.

16. Li, L. et al. Development of a direct reverse-transcription quantitative PCR (dirRT-qPCR) assay for clinical Zika diagnosis. International Journal of Infectious Diseases 85, 167–174 (2019).

17. Smyrlaki, I. et al. Massive and rapid COVID-19 testing is feasible by extraction-free SARS-CoV-2 RT-qPCR. *medRxiv* 2020.04.17.20067348 (2020) doi:10.1101/2020.04.17.20067348.

18. Bhadra, S., Riedel, T. E., Lakhotia, S., Tran, N. D. & Ellington, A. D. High-surety isothermal amplification and detection of SARS-CoV-2, including with crude enzymes. *bioRxiv* 2020.04.13.039941 (2020) doi:10.1101/2020.04.13.039941.

19. Bhadra, S., Saldaña, M. A., Han, H. G., Hughes, G. L. & Ellington, A. D. Simultaneous Detection of Different Zika Virus Lineages via Molecular Computation in a Point-of-Care Assay. Viruses 10, 714 (2018).

20. Wang, R., Hozumi, Y., Yin, C. & Wei, G.-W. Mutations on COVID-19 diagnostic targets. *arXiv:2005.02188 [q-bio]* (2020).

21. Larremore, D. B. et al. Test sensitivity is secondary to frequency and turnaround time for COVID-19 surveillance. *medRxiv* 2020.06.22.20136309 (2020) doi:10.1101/2020.06.22.20136309.

22. He, X. et al. Temporal dynamics in viral shedding and transmissibility of COVID-19. Nature Medicine 1-4 (2020) doi:10.1038/s41591-020-0869-5.

23. Oscorbin, I. P., Belousova, E. A., Zakabunin, A. I., Boyarskikh, U. A. & Filipenko, M. L. Comparison of fluorescent intercalating dyes for quantitative loop-mediated isothermal amplification (qLAMP). BioTechniques 61, 20–25 (2016).

24. Zhang, Y. et al. Enhancing Colorimetric LAMP Amplification Speed and Sensitivity with Guanidine Chloride. *bioRxiv* 2020.06.03.132894 (2020) doi:10.1101/2020.06.03.132894.

25. Tomita, N., Mori, Y., Kanda, H. & Notomi, T. Loop-mediated isothermal amplification (LAMP) of gene sequences and simple visual detection of products. Nature Protocols 3, 877–882 (2008).

26. Hsieh, K., Mage, P. L., Csordas, A. T., Eisenstein, M. & Soh, H. T. Simultaneous elimination of carryover contamination and detection of DNA with uracil-DNA-glycosylase-supplemented loop-mediated isothermal amplification (UDG-LAMP). Chem. Commun. 50, 3747–3749 (2014).

27. Wang, X. et al. Rapid and sensitive detection of Zika virus by reverse transcription loop-mediated isothermal amplification. Journal of Virological Methods 238, 86–93 (2016).

28. Osterdahl, M. F. et al. Detecting SARS-CoV-2 at point of care: Preliminary data comparing Loop-mediated isothermal amplification (LAMP) to PCR. *medRxiv* 2020.04.01.20047357 (2020) doi:10.1101/2020.04.01.20047357.

29. Park, G.-S. et al. Development of Reverse Transcription Loop-mediated Isothermal Amplification (RT-LAMP) Assays Targeting SARS-CoV-2. *bioRxiv* 2020.03.09.983064 (2020) doi:10.1101/2020.03.09.983064.

30. Schmid-Burgk, J. L. et al. LAMP-Seq: Population-Scale COVID-19 Diagnostics Using a Compressed Barcode Space. *bioRxiv* 2020.04.06.025635 (2020) doi:10.1101/2020.04.06.025635.

31. Butler, D. J. et al. Host, Viral, and Environmental Transcriptome Profiles of the Severe Acute Respiratory Syndrome Coronavirus 2 (SARS-CoV-2). http://biorxiv.org/lookup/doi/10.1101/2020.04.20.048066 (2020) doi:10.1101/2020.04.20.048066.

32. Ben-Assa, N. et al. SARS-CoV-2 On-the-Spot Virus Detection Directly From Patients. *medRxiv* 2020.04.22.20072389 (2020) doi:10.1101/2020.04.22.20072389.

33. Srivatsan, S. et al. Preliminary support for a ‘dry swab, extraction free’ protocol for SARS-CoV-2 testing via RT-qPCR. bioRxiv 2020.04.22.056283 (2020) doi:10.1101/2020.04.22.056283.

34. Tu, Y.-P. et al. Patient-collected tongue, nasal, and mid-turbinate swabs for SARS-CoV-2 yield equivalent sensitivity to health care worker collected nasopharyngeal swabs. *medRxiv* 2020.04.01.20050005 (2020) doi:10.1101/2020.04.01.20050005.

35. Wang, W. et al. Detection of SARS-CoV-2 in Different Types of Clinical Specimens. JAMA (2020) doi:10.1001/jama.2020.3786.

36. Ochert, A. S., Boulter, A. W., Birnbaum, W., Johnson, N. W. & Teo, C. G. Inhibitory effect of salivary fluids on PCR: potency and removal. Genome Res. 3, 365–368 (1994).

37. Wei, S. et al. Field-deployable, rapid diagnostic testing of saliva samples for SARS-CoV-2. *medRxiv* 2020.06.13.20129841 (2020) doi:10.1101/2020.06.13.20129841.

38. R Core Team. R: A Language and Environment for Statistical Computing. (R Foundation for Statistical Computing, 2018).

39. Wickham, H. ggplot2: Elegant Graphics for Data Analysis. (Springer-Verlag New York, 2016).

40. Pastorino, B., Touret, F., Gilles, M., Lamballerie, X. de & Charrel, R. N. Evaluation of heating and chemical protocols for inactivating SARS-CoV-2. *bioRxiv* 2020.04.11.036855 (2020) doi:10.1101/2020.04.11.036855.

